# Outcome of Conservative Therapy in COVID-19 Patients Presenting with Gastrointestinal Bleeding

**DOI:** 10.1101/2020.08.06.20169813

**Authors:** DM Shalimar, Manas Vaishnav, Anshuman Elhence, Ramesh Kumar, Srikant Mohta, Chandan Palle, Peeyush Kumar, Mukesh Ranjan, Tanmay Vajpai, Shubham Prasad, Jatin Yegurla, Anugrah Dhooria, Vikas Banyal, Samagra Agarwal, Rajat Bansal, Sulagna Bhattacharjee, Richa Aggarwal, Kapil Dev Soni, Swetha Rudravaram, Ashutosh Kumar Singh, Irfan Altaf, Avinash Choudekar, Soumya Jagannath Mahapatra, Deepak Gunjan, Saurabh Kedia, Govind Makharia, Anjan Trikha, Pramod Garg, Anoop Saraya

## Abstract

**Background/Objective:** There is a paucity of data on the management of gastrointestinal (GI) bleeding in patients with COVID-19 amid concerns about the risk of transmission during endoscopic procedures. We aimed to study the outcomes of conservative treatment for GI bleeding in patients with COVID-19.

**Methods:** In this retrospective analysis, 24 of 1342 (1.8%) patients with COVID-19, presenting with GI bleeding from 22 April to 22 July 2020, were included.

**Results:** The mean age of patients was 45.8±12.7 years; 17 (70.8%) were males; upper GI (UGI) bleeding: lower GI (LGI) 23:1. Twenty-two (91.6%) patients had evidence of cirrhosis-21 presented with UGI bleeding while one had bleeding from hemorrhoids. Two patients without cirrhosis were presumed to have non-variceal bleeding. The medical therapy for UGI bleeding included vasoconstrictors-somatostatin in 17 (73.9%) and terlipressin in 4 (17.4%) patients. All patients with UGI bleeding received proton pump inhibitors and antibiotics. Packed red blood cells (PRBCs), fresh frozen plasma and platelets were transfused in 14 (60.9%), 3 (13.0%) and 3 (13.0%), respectively. The median PRBCs transfused was 1 (0-3) unit(s). The initial control of UGI bleeding was achieved in all 23 patients and none required an emergency endoscopy. At 5-day follow-up, none rebled or died. Two patients later rebled, one had intermittent bleed due to gastric antral vascular ectasia, while another had rebleed 19 days after discharge. Three (12.5%) cirrhosis patients succumbed to acute hypoxemic respiratory failure during hospital stay.

**Conclusion:** Conservative management strategies including pharmacotherapy, restrictive transfusion strategy, and close hemodynamic monitoring can successfully manage GI bleeding in COVID-19 patients and reduce need for urgent endoscopy. The decision for proceeding with endoscopy should be taken by a multidisciplinary team after consideration of the patient’s condition, response to treatment, resources and the risks involved, on a case to case basis.

## Introduction

Coronavirus disease 2019 (COVID-19) cases continue to rise and have exceeded 15 million worldwide to date. Recent data suggest that the overall prevalence of chronic liver disease (CLD) among COVID-19 patients is around 3-4%.[1] Patients with CLD and cirrhosis are at higher risk of mortality from COVID-19, especially those with acute-on-chronic liver failure.[1,2] The clinical presentation of COVID-19 in patients with CLD and cirrhosis, apart from respiratory symptoms, may include acute decompensation (AD) with gastrointestinal (GI) bleeding.[1]

The severe acute respiratory syndrome coronavirus-2 (SARS CoV-2) spreads via droplet and fomites.[3] The presence of a viable virus in droplet nuclei <5 µm, which can travel a longer distance, has been demonstrated in artificially generated experiments.[4,5] Upper GI endoscopy is an aerosol-generating procedure with the antecedent risk of transmission of the virus to the endoscopist as well as the assistant. The risks associated with transmission during endoscopy has led to many expert societies advocating for avoidance of elective endoscopic procedures.[6-9]

GI bleeding as a presentation in patients with COVID-19 is an emergency. The associated CLD, coagulopathy, use of anticoagulants, mechanical ventilation and comorbidities may predispose COVID-19 patients to GI bleeding.[10] Upper GI (UGI) bleeding is associated with high 42-day mortality in patients with cirrhosis.[11] The risks associated with the transmission of COVID-19 have led to a dilemma in the management of such patients. Most society guidelines based on expert consensus recommend that emergency endoscopies can be done.[6-9] There is a paucity of data on the management and outcomes of UGI bleeding in patients with COVID-19. We aimed to assess the clinical presentations and outcomes of COVID-19 patients admitted with GI bleeding.

## Methods

### Patient population

In this retrospective study, consecutive COVID-19 patients with GI bleed who presented to the Gastroenterology department, or dedicated COVID-19 care facility of All India Institute of Medical Sciences (AIIMS), New Delhi, India between 22 April 2020 and 22 July 2020 were included. Patients were diagnosed as COVID-19 based on reverse transcriptase-polymerase chain reaction (RT-PCR) positivity, as per the Indian Council of Medical Research (ICMR) criteria. Data was collected from a prospectively maintained database of all patients from admission until the endpoint of the study (discharge or death or deadline of 22 July 2020).

### Patient evaluation and management

All patients presenting with GI bleeding to the emergency with either fever or respiratory symptoms or chest X-ray showing infiltrates were tested for COVID-19 as per the hospital policy. Nasal and throat swabs were taken and transported in a viral transport media (Hank’s balanced salt solution) and tested by reverse transcriptase-polymerase chain reaction (RT-PCR).

Cirrhosis was defined by the presence of a nodular outline of liver on imaging and concomitant evidence of portal hypertension, such as a dilated portal vein and splenomegaly. UGI bleeding was defined as bleeding originating from the esophagus up to the ligament of Treitz at the duodenojejunal junction, while lower gastrointestinal (LGI) bleeding was defined as bleeding originating from distal to the ligament of Treitz.[12] In the absence of endoscopic evidence, those presenting with a background of cirrhosis with either hematemesis with melena or blood in nasogastric tube at presentation were classified as having UGI bleeding. Those presenting with fresh bleeding per rectum in absence of blood in the nasogastric tube were classified as LGI bleeding.

In absence of endoscopy, control of bleeding was defined as absence of fresh blood in the nasogastric tube, clearing of melena, no further drop in hemoglobin levels, and absence of tachycardia or hypotension.

All patients were managed according to standard guidelines, as feasible.[13] Briefly, the patients were resuscitated at presentation and started on a splanchnic vasoconstrictor drug (either terlipressin or somatostatin, for a duration of 3-5 days), if there was evidence of cirrhosis or portal hypertension, either clinically or on imaging. Patients were given packed red blood cells (PRBC) to keep a target hemoglobin of 7 to 8 g/dL. Proton pump inhibitor (pantoprazole) was given to all patients either as an infusion or twice-daily injections. All patients were given antibiotics as primary prophylaxis. All patients underwent close hemodynamic monitoring for any evidence of persistent bleeding, as defined above. Monitoring of hemoglobin, heart rate, blood pressure and any fresh blood in the nasogastric tube was done.

Endoscopy was considered as a therapeutic modality if there was a suspicion of active bleeding after 24 hours.

The Glasgow-Blatchford bleeding score (GBS), modified Glasgow-Blatchford bleeding score (mGBS), clinical Rockall score (CRS), and albumin, international normalized ratio, mental status, systolic blood pressure and age >65 (AIMS65) score were calculated as defined.[14-17]

Carvedilol was started as secondary prophylaxis (on day 3-5) for the patients with suspected variceal bleeding, after stopping of vasoactive drugs and when patient was hemodynamically stable. The hospital stay was counted onwards from the date of COVID-19 detection for cases diagnosed during their hospitalization and from the date of admission for those who presented with a positive test.

### Management of COVID-19

The respiratory clinical presentations included cough, breathlessness and pneumonia. The clinical severity of COVID-19 was defined according to the Ministry of Health and Family Welfare (MOHFW) criteria as follows-mild disease as patients with only upper respiratory tract symptoms without any signs of breathlessness and hypoxia. Moderate severity was defined as the presence of pneumonia with the respiratory rate (RR) between 24-30/minute and SpO_2_ between 90-94% on room air, while severe disease was defined by the presence of pneumonia with RR >30/minute or SpO_2_ <90% on room air or severe respiratory distress.

The specific therapies for the management of COVID-19 included hydroxychloroquine and Azithromycin. All patients received supplemental vitamin C, and Zinc. Oxygen and ventilatory support were used, as per the evolving consensus and at the discretion of the treating physician, in the best interest of the patient.

### Statistical analysis

Normally distributed data were expressed as mean ± standard deviation, and skewed data were expressed as median with interquartile range. The normality of the data was tested with Shapiro-Wilk test, p-value >0.05 indicated normal distribution and <0.05 indicated skewed data. Nominal data were expressed as frequency or percentages. Data were analyzed using IBM SPSS statistical software (Version 20.0, Chicago, IL, USA).

### Ethical clearance

In this retrospective study, the requirement of consent was waived off, and ethical approval was obtained from the Institutional Ethics Committee, All India Institute of Medical Sciences, India (Ref No: IEC-253/17.04.2020).

## Results

Twenty four of 1342 (1.8%) COVID-19 patients admitted during the study period had GI bleeding. The mean age of the patients was 45.8 ± 12.7 years and 17 (70.8%) were males. The clinical presentation included hematemesis alone in 12 (50%), melena alone in 4 (16.7%) and combined hematemesis and melena in 7 (29.1%) patients, whereas only one (4.2%) patient had fresh bleeding per rectum (hemorrhoidal bleeding). At presentation, the median hemoglobin was 7.2 (5.8-9.0) g/dL, platelet count (x10^3^/mm^3^) was 90.5 (52-135) and international normalized ratio (INR) was 1.2 (1.2-1.40) (Table 1). Of the 24 patients, 22 (91.7%) had CLD.

**Table 1.**
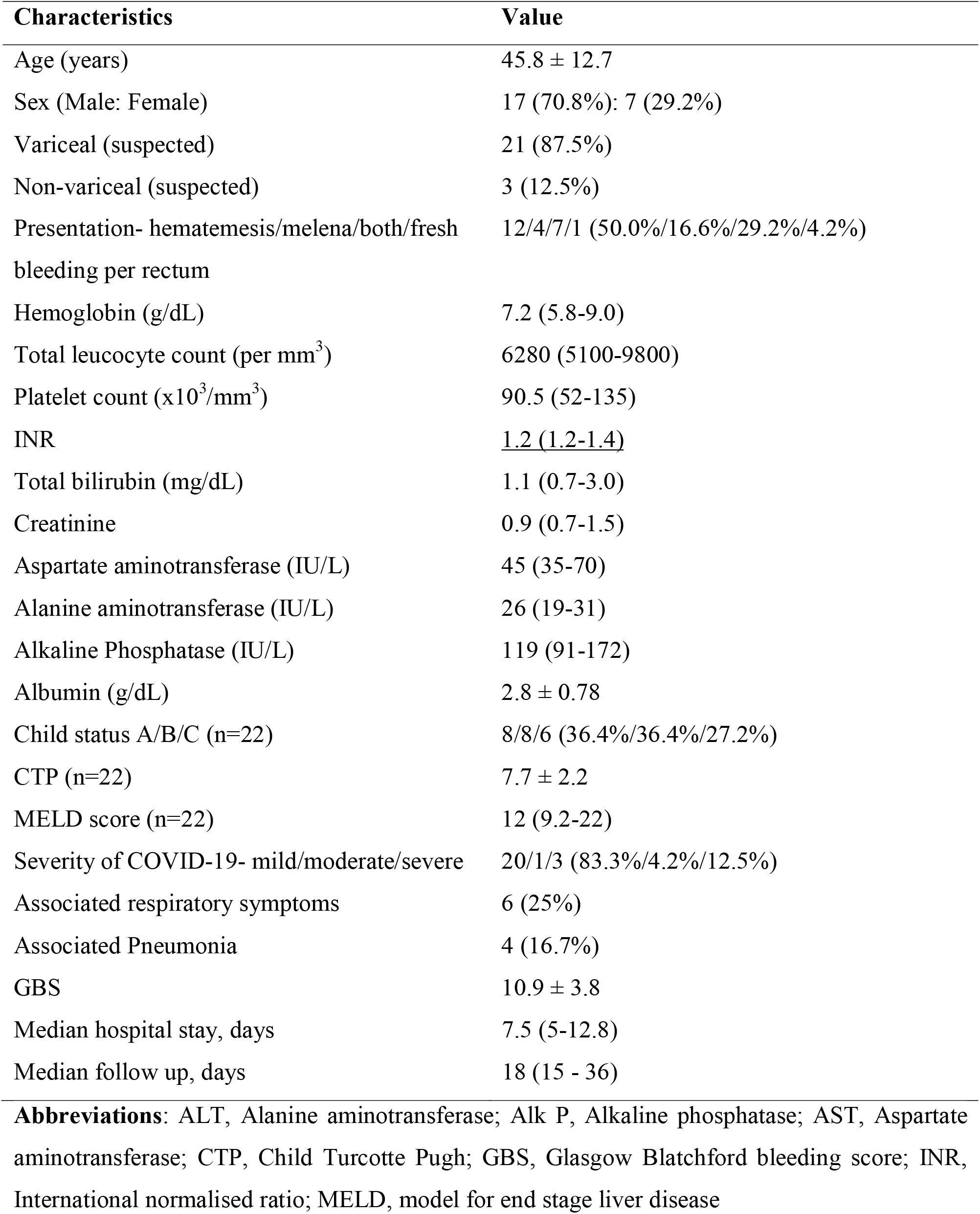
Demographics of all patients with bleeding admitted during the study period (n=24).

Among the 22 patients with suspected CLD, the median model for end-stage liver disease (MELD) score was 12 (9.7-22), and Child-Pugh-Turcotte class A/B/C were 8/8/6 patients, respectively. At presentation, in the absence of endoscopy, we suspected portal hypertension-related bleeding in 21 (87.5%) patients. The etiology of CLD included alcohol 8 (36.4%), cryptogenic 6 (27.3%), viral 4 (18.2%), autoimmune hepatitis (AIH) and other etiologies in 2 (9.1%) each, respectively.

Of the 3 (12.5%) suspected non-portal hypertension-related bleeds, one patient had CLD with hemorrhoidal bleed, and two patients presented with hematemesis and melena without any evidence of underlying CLD. Amongst these 2 patients, one was on dual antiplatelets for secondary prophylaxis of cardiovascular disease, and in the other patient, no obvious cause for GI bleed could be attributed.

The severity of COVID-19 was mild, moderate and severe in 20 (83.3%), 1 (4.2%) and 3 (12.5%) patients, respectively. Six (25%) patients had associated respiratory symptoms, and 4 (16.7%) patients developed pneumonia. The median hospital stay was 7.5 (5-12.75) days.

### Risk factors for UGI bleed

Past history of variceal bleed was present in 14/23 (60.9%). History of hepatic encephalopathy was present in 3 (13.0%), platelet count (<50000/mm^3^) in 5 (21.7%), INR (>2) in 3 (13.0%), and creatinine (>1 mg/dl) in 9 (39.1%). Only one patient had Child C and creatinine (>1 mg/dl). The median GBS, mGBS and CRS were 12 (9-14), 10 (7-11) and 3 (3-4), respectively (Table 2).

**Table 2.** Management of UGI bleeding (n=23).

| **Drugs** | **N (%)** |
| --- | --- |
| Somatostatin | 17 (73.9%) |
| Terlipressin | 4 (17.4%) |
| PPI | 23 (100%) |
| FFP | 3 (13.0%) |
| Vit K | 9 (39.1%) |
| Antifibrinolytics | 2 (8.7%) |
| Blood transfusion (PRBCs) | 14 (60.9%) |

**Abbreviations:** FFP, Fresh frozen plasma; PPI, Proton pump inhibitors; PRBC, Packed red blood cells; Vit, Vitamin.

### Management of UGI bleed

None of the patients had shock or fresh blood in the nasogastric tube at presentation. Vasoactive agents, somatostatin in 17 (73.9%) and terlipressin in 4 (17.4%), were used in patients with suspected portal hypertension related bleeding (Table 3). Proton pump inhibitors were used in all 23 (100%) patients with UGI bleeding. PRBCs transfusions were given in 14 (60.9%) patients. The median PRBCs transfused was 1 (0-3) unit(s). FFP transfusion and platelet transfusion were given in 3 (13.0%) patients each, respectively. Vitamin K was given in 9 (39.1%), and 2 (8.7%) patients received antifibrinolytics.

**Table 3.** Risk factors for UGI bleeding (n=23)

| **Risk factor** | **N (%)** |
| --- | --- |
| Hypotension /shock | 0 |
| Active bleeding in nasogastric tube | 0 |
| Past history of variceal bleeding | 14 (60.9%) |
| GBS | 12 (9-14) |
| mGBS | 10 (7-11) |
| CRS | 3 (3-4) |
| AIMS65 | 1 (0-1) |
| Hepatic encephalopathy | 3 (13.0%) |
| INR (>2) | 3 (13.0%) |
| Creatinine (> 1 mg/dl) | 9 (39.1%) |
| Platelet (<50000) | 5 (21.7%) |

**Abbreviations:** AIMS65, albumin, international normalised ratio, mental status, systolic blood pressure age>65; CRS, clinical Rockall score; GBS, Glasgow-Blatchford bleeding score; INR, international normalised ratio; mGBS, modified Glasgow-Blatchford bleeding score.

### Outcomes of UGI bleed

With above mentioned conservative treatment, the UGI bleeding could be controlled in all 23 (100%) patients within 24 hours, and none required an emergency endoscopy. None of the patients rebled or died at 5 days (Table 4). During hospitalization, 3 patients with cirrhosis died after 7, 12 and 20 days of admission. All these 3 patients had associated pneumonia and required mechanical ventilation. The cause of death in all 3 patients was acute hypoxemic respiratory failure followed by multi-organ failure. There was no active bleeding in these 3 patients.

**Table 4.** Outcomes of patients with UGI bleeding (n=23).

| Outcome | N (%) |
| --- | --- |
| Rebleed at 5 days | 0 |
| Mortality at 5 days | 0 |
| In-hospital mortality | 3 (13.0%) |
| Rebleeding at follow-up | 2 (8.7%) |
| Mortality at last follow-up | 3 (13.0%) |

Out of the remaining 20 patients, follow up data was available in 17 (85%) patients, and 3 (15%) were lost to follow up. The median follow-up duration was 18 (15-36) days. One patient with gastric antral vascular ectasia (GAVE) continued to bleed intermittently and underwent UGI endoscopy with argon plasma coagulation application after 18 days of initial admission. Another patient had rebleeding on the 19^th^ day after hospital discharge lasting for 2 days and resolved spontaneously; presently, the patient is on carvedilol. No mortality was reported post-discharge during follow up. Three patients complained of increasing ascites. None developed hepatic encephalopathy.

### LGI bleed

One patient presented with fresh bleeding per rectum and with a background of interstitial lung disease and CLD. An evaluation revealed internal hemorrhoids with active bleeding. The patient was managed with rectal packing. No PRBCs were transfused. There was no rebleed after removing the rectal packing.

## Discussion

The current COVID-19 pandemic has changed the paradigms of management of patients with GI bleeding. The risk associated with the transmission of COVID-19 during endoscopic procedures has led to a more conservative approach in the management of such patients. In our study, majority of patients who presented with upper GI bleeding were clinically suspected to be related to portal hypertension. They responded well to conservative management and hence emergency endoscopic treatment could be avoided. The emphasis should be on resuscitation and rapid institution of pharmacological therapy with splanchnic vasoconstrictors, proton pump inhibitors and antibiotics.

It is well known that nearly 50% of variceal bleeds stop spontaneously without any treatment.[18] Baveno VI guidelines recommend vasoconstrictors in combination with endoscopic therapy to decrease the rebleeding rates.[13] Lo et al. compared terlipressin alone vs terlipressin plus band ligation in patients with no active variceal bleeding on endoscopy and reported a higher 5-day treatment failure (15% vs 0%, p=0.006) for terlipressin alone group. However, both groups had similar 6-week mortality and complication rates. It is important to note that 48 hour bleeding control was more than 90% in both groups.[19] There is also a single trial comparing somatostatin alone vs combination therapy with somatostatin and endoscopic therapy, which showed higher failure to control acute bleeding in somatostatin alone (24% vs 8%; p=0.03), but the 6-week mortality was equal in both groups, and on multivariate analysis, presence of shock, and active bleeding at endoscopy predicted the failure of somatostatin.[20]

With our management protocol, we achieved control of bleeding in all except one patient, and none of the patients had active bleeding and mortality at 5 days. The 3 patients who died during hospital stay had developed acute hypoxemic respiratory failure followed by multi-organ failure, likely because of COVID-19 pneumonia. There was no ongoing active bleeding in them. However, it is possible that GI bleed may have triggered the worsening in these patients. In patients with cirrhosis and variceal bleeding, the reported failure rate to control bleeding at 5-days varies from 10-to 20%, and mortality at 5-days is 2-5%.[21] The 42-days mortality varies from 12-20%.[22-24] The rebleeding and mortality in these patients is determined by multiple factors, most importantly by underlying liver disease status. Only one patient in our study group had Child-Pugh C cirrhosis with creatinine (>1 mg/dl) and three had developed HE, while the median MELD score was 12 and none had shock, hepatocellular carcinoma, which are the factors associated with poor outcome.[25,26] COVID-19 pneumonia was severe in only 3 patients. These facts suggest that most of the patients included in this study did not have poor prognostic factors. We used terlipressin or somatostatin as vasoactive agents. A recent meta-analysis reported no difference in rebleeding and mortality with either of these drugs.[27] All patients with suspected variceal bleed received carvedilol as secondary prophylaxis. Carvedilol has been shown to decrease mortality and reduce hepatic venous pressure gradient (HVPG) more than other non-selective beta-blockers.[28,29] Of all the patients included in the present study, outcomes of 13 patients have been described in our previous publication.[1]

A recent study from the United States reported outcomes of patients with COVID-19 pneumonia and GI bleed managed conservatively.[10] This study included 6 patients with likely ulcer-related bleeding and mean GBS of 13.5. All patients responded to conservative management, and none required endoscopy. The authors proposed a lack of response at the end of 24 hours to be an indication for endoscopy.[10] Our results validate these findings in a study population of predominantly CLD. A multicenter study from New York, USA suggested that patients admitted with GI bleeding during the COVID-19 pandemic were more likely to have a longer hospital stay (OR 2.62), transfusion requirement (OR 2.86) and lesser odds of undergoing an endoscopy (OR 0.32) as compared to the pre-COVID era.[30] In our study, the median hospital stay was 7.5 days and 60% of patients received PRBCs or other fresh frozen plasma. We were conservative in the transfusion of PRBCs, with a targeted hemoglobin of 7-8 g/dl. Prior studies suggest that excessive transfusion of blood products and FFP/platelet transfusion are associated with increased risk of bleeding.[31]

Overall, 90% of all patients in our study were suspected of having portal hypertension-related bleeding. However, it is quite possible that some of them could have non-variceal bleeding. We managed patients with standard pharmacological therapy, including a combination of splanchnic vasoconstrictors and other drugs like proton pump inhibitors and vitamin K, with successful response. All patients were monitored by a multidisciplinary panel comprising gastroenterologists, emergency physicians and anesthesiologists. The clinical decisions were based on the assessment of active bleeding-the presence of fresh blood in nasogastric tube among patients with hematemesis, drop in hemoglobin, and new-onset tachycardia. We did not consider endoscopy at the initial presentation but kept it as a back-up for actively bleeding patients. Only one patient required an endoscopy after 18 days of admission for intermittent bleeding due to GAVE and was successfully managed with APC application. The Baveno VI guidelines recommend endoscopy within 12 hours of presentation in patients with UGI bleeding and features of cirrhosis.[13] Delaying endoscopy up to 24 hours has not been shown to affect 30-day mortality in a recent study.[32] The performance of endoscopy in patients with COVID-19 depends on multiple challenging factors, including the indication for the procedure, availability of personal protective equipment (PPE), trained personnel, and appropriate infrastructure. All major societies recommend rescheduling non-urgent endoscopic procedures and performing endoscopy in COVID positive patients or those awaiting results with isolation precautions in negative pressure rooms.[6-9] In the absence of negative pressure rooms in our existing infrastructure, we did make arrangements for emergency endoscopies, in case needed.

The prognostic scores such as CRS, GBS, mGBS and AIMS65 predict the need for hospitalization, rebleeding and mortality better among patients with non-variceal UGI bleeding than patients with variceal UGI bleeding.[22] The majority of patients in our study had variceal UGI bleed, and despite having unfavorable prognostic scores, the bleeding was controlled in all. Overall, the incidence of GI bleeding in COVID patients without CLD was extremely low, 2/1320 (0.2%). This could be related to no upper or lower GI involvement by the virus to the extent of causing mucosal inflammation, younger patients, and lack of NSAIDs use.

Our study has a few limitations. Small sample size, single-centre retrospective design with predominantly descriptive nature of the study are its inherent limitations. In the absence of endoscopy, we were not sure of the exact cause of UGI bleeding in all patients. We presumed all patients with cirrhosis to have portal hypertension-related bleeding. It is well known that up to 25% of patients with cirrhosis may have a non-variceal cause of bleeding. The inclusion of patients predominantly with CLD limits generalizability to other causes of bleed. We had only one patient with a LGI bleed.

In conclusion, conservative management strategies including pharmacotherapy, restrictive transfusion strategy, and close hemodynamic monitoring can successfully manage GI bleeding in COVID-19 patients and reduce the need for urgent endoscopy. The decision for proceeding with endoscopy should be taken by a multidisciplinary team after consideration of the patient’s condition, response to treatment, resources and the risks involved, on a case to case basis.

## Data Availability

All data regarding the study is available with the corresponding author.

## Declaration of interests

We declare no competing interests

## Disclosures

None for all authors

All authors approved the final manuscript

## Funding source

None

## Acknowledgements

The study was supported by a grant from All India Institute of Medical Sciences, New Delhi. Grant number A-COVID-28; 02.06.2020.

We like to thank the Clinical Research Unit, All India Institute of Medical Sciences, New Delhi.

We like to thank the Department of Emergency Medicine, Department of Anesthesiology, Pain medicine and Critical Care and Department of Laboratory Medicine, All India Institute of Medical Sciences, New Delhi.

We like to thank Indian Council of Medical Research (ICMR) for providing kits for SARS-CoV2 testing.

Conflict on interest: None for all authors

## Notes

### Competing Interest Statement

The authors have declared no competing interest.

### Funding Statement

The study was supported by AIIMS Intramural grant

### Author Declarations

Institutional Ethics Committee, All India Institute of Medical Sciences, India (Ref No: IEC-253/17.04.2020)

## References

1. Shalimar, Elhence A, Vaishnav M, et al. Poor Outcomes in Patients with Cirrhosis and COVID-19. Indian J Gastroenterol.2020;39. doi.org/10.1007/s12664-020-01074-3

2. Iavarone M, D’Ambrosio R, Soria A, et al. High rates of 30-day mortality in patients with cirrhosis and COVID-19. J Hepatol. 2020;

3. Transmission of SARS-CoV-2: implications for infection prevention precautions [Internet]. [cited 2020 Jul 26]. Available from: https://www.who.int/news-room/commentaries/detail/transmission-of-sars-cov-2-implications-for-infection-prevention-precautions

4. van Doremalen N, Bushmaker T, Morris DH, et al. Aerosol and Surface Stability of SARS-CoV-2 as Compared with SARS-CoV-1. N Engl J Med. 2020;382:1564–7.

5. Anfinrud P, Stadnytskyi V, Bax CE, Bax A. Visualizing Speech-Generated Oral Fluid Droplets with Laser Light Scattering. N Engl J Med. 2020;382:2061–3.

6. Sawhney MS, Bilal M, Pohl H, et al. Triaging advanced GI endoscopy procedures during the COVID-19 pandemic: consensus recommendations using the Delphi method. Gastrointest Endosc. 2020;

7. Philip M, Lakhtakia S, Aggarwal R, Madan K, Saraswat V, Makharia G. Joint Guidance from SGEI, ISG and INASL for Gastroenterologists and Gastrointestinal Endoscopists on the Prevention, Care, and Management of Patients With COVID-19. J Clin Exp Hepatol. 2020;10:266–70.

8. Gralnek IM, Hassan C, Beilenhoff U, et al. ESGE and ESGENA Position Statement on gastrointestinal endoscopy and the COVID-19 pandemic. Endoscopy. 2020;52:483–90.

9. Joint GI society message: COVID-19 clinical insights for our community of gastroenterologists and gastroenterology care providers | American Gastroenterological Association [Internet]. [cited 2020 Jul 26]. Available from: https://gastro.org/press-releases/joint-gi-society-message-covid-19-clinical-insights-for-our-community-of-gastroenterologists-and-gastroenterology-care-providers/

10. Cavaliere K, Levine C, Wander P, Sejpal DV, Trindade AJ. Management of upper GI bleeding in patients with COVID-19 pneumonia. Gastrointest Endosc. 2020;92:454–5.

11. Rout G, Shalimar, Gunjan D, et al. Thromboelastography-guided Blood Product Transfusion in Cirrhosis Patients With Variceal Bleeding: A Randomized Controlled Trial. J Clin Gastroenterol. 2020;54:255–62.

12. Kim BSM, Li BT, Engel A, et al. Diagnosis of gastrointestinal bleeding: A practical guide for clinicians. World J Gastrointest Pathophysiol. 2014;5:467–78.

13. Franchis R de. Expanding consensus in portal hypertension: Report of the Baveno VI Consensus Workshop: Stratifying risk and individualizing care for portal hypertension. J Hepatol. 2015;63:743–52.

14. Blatchford O, Murray WR, Blatchford M. A risk score to predict need for treatment for upper-gastrointestinal haemorrhage. Lancet. 2000;356:1318–21.

15. Saltzman JR, Tabak YP, Hyett BH, Sun X, Travis AC, Johannes RS. A simple risk score accurately predicts in-hospital mortality, length of stay, and cost in acute upper GI bleeding. Gastrointest Endosc. 2011;74:1215–24.

16. Rockall TA, Logan RF, Devlin HB, Northfield TC. Risk assessment after acute upper gastrointestinal haemorrhage. Gut. 1996;38:316–21.

17. Romagnuolo J, Barkun AN, Enns R, Armstrong D, Gregor J. Simple clinical predictors may obviate urgent endoscopy in selected patients with nonvariceal upper gastrointestinal tract bleeding. Arch Intern Med. 2007;167:265–70.

18. D’Amico G, Pagliaro L, Bosch J. Pharmacological treatment of portal hypertension: an evidence-based approach. Semin Liver Dis. 1999;19:475–505.

19. Lo G-H, Chen W-C, Wang H-M, et al. Low-dose terlipressin plus banding ligation versus low-dose terlipressin alone in the prevention of very early rebleeding of oesophageal varices. Gut. 2009;58:1275–80.

20. Villanueva C, Ortiz J, Sàbat M, et al. Somatostatin alone or combined with emergency sclerotherapy in the treatment of acute esophageal variceal bleeding: a prospective randomized trial. Hepatology. 1999;30:384–9.

21. D’Amico G, De Franchis R, Cooperative Study Group. Upper digestive bleeding in cirrhosis. Post-therapeutic outcome and prognostic indicators. Hepatology. 2003;38:599–612.

22. Rout G, Sharma S, Gunjan D, Kedia S, Nayak B, Shalimar. Comparison of various prognostic scores in variceal and non-variceal upper gastrointestinal bleeding: A prospective cohort study. Indian J Gastroenterology. 2019;38:158–66.

23. Bambha K, Kim WR, Pedersen R, Bida JP, Kremers WK, Kamath PS. Predictors of early rebleeding and mortality after acute variceal haemorrhage in patients with cirrhosis. Gut. 2008;57:814–20.

24. Altamirano J, Zapata L, Agustin S, et al. Predicting 6-week mortality after acute variceal bleeding: role of Classification and Regression Tree analysis. Ann Hepatol. 2009;8:308–15.

25. Malinchoc M, Kamath PS, Gordon FD, Peine CJ, Rank J, ter Borg PC. A model to predict poor survival in patients undergoing transjugular intrahepatic portosystemic shunts. Hepatology. 2000;31:864–71.

26. Rout G, Sharma S, Gunjan D, et al. Development and Validation of a Novel Model for Outcomes in Patients with Cirrhosis and Acute Variceal Bleeding. Dig Dis Sci. 2019;64:2327-37.

27. Zhou X, Tripathi D, Song T, et al. Terlipressin for the treatment of acute variceal bleeding. Medicine (Baltimore). 2018;97(48):e13427

28. Gupta V, Rawat R, Shalimar, Saraya A. Carvedilol versus propranolol effect on hepatic venous pressure gradient at 1 month in patients with index variceal bleed: RCT. Hepatol Int. 2017;11:181–7.

29. Sinha R, Lockman KA, Mallawaarachchi N, Robertson M, Plevris JN, Hayes PC. Carvedilol use is associated with improved survival in patients with liver cirrhosis and ascites. J Hepatol. 2017;67:40–6.

30. Kim J, Doyle JB, Blackett JW, et al. Effect of the COVID-19 Pandemic on Outcomes for Patients Admitted with Gastrointestinal Bleeding in New York City. Gastroenterology. 2020;

31. Rockey DC. To transfuse or not to transfuse in upper gastrointestinal hemorrhage? That is the question. Hepatology. 2014;60:422–4.

32. Lau JYW, Yu Y, Tang RSY, et al. Timing of Endoscopy for Acute Upper Gastrointestinal Bleeding. N Engl J Med. 2020;382:1299–308.

